# The asymptomatic proportion of SARS-CoV-2 Omicron-variant infections in households: A systematic review

**DOI:** 10.1101/2024.04.15.24305816

**Authors:** Nancy DJ Shi, Adrian J Marcato, Violeta Spirkoska, Niamh Meagher, Juan-Pablo Villanueva-Cabezas, David J Price

## Abstract

**Background:** Understanding the clinical spectrum of SARS-CoV-2 infection, including the asymptomatic fraction, is important as asymptomatic individuals are still able to infect other individuals and contribute to ongoing transmission. The WHO Unity Household transmission investigation (HHTI) protocol provides a platform for the prospective and systematic collection of high-quality clinical, epidemiological, serological, and virological data from SARS-CoV-2 confirmed cases and their household contacts. These data can be used to understand key severity and transmissibility parameters — including the asymptomatic proportion — in relation to local epidemic context and help inform public health response.

**Methods:** We aimed to estimate the asymptomatic proportion of SARS-CoV-2 Omicron-variant infections in Unity-aligned HHTIs. We conducted a systematic review and meta-analysis in alignment with the PRISMA 2020 guidelines and registered our systematic review on PROSPERO (CRD42022378648). We searched EMBASE, Web of Science, MEDLINE, and bioRxiv and medRxiv from 1 November 2021 to 22 August 2023.

**Results:** We identified 8,368 records, of which 98 underwent full text review. We identified only three studies for data extraction, with substantial variation in study design and corresponding estimates of the asymptomatic proportion. As a result, we did not generate a pooled estimate or *I*^*2*^ metric.

**Conclusions:** The limited number of quality studies that we identified highlights the need for improved preparedness and response capabilities to facilitate robust HHTI implementation, analysis and reporting, to better inform national, regional and global risk assessments and policy making.

**Key messages:**

- Estimates for the asymptomatic proportion of SARS-CoV-2 Omicron-variant infections are highly heterogeneous
- We assessed the proportion of SARS-CoV-2 Omicron-variant infections among household contacts, who were followed prospectively and systematically, per the WHO Unity household transmission investigation protocol.
- Given the small number of studies with sufficient data and the observed heterogeneity in the asymptomatic proportion point estimates, we did not provide a pooled estimate of the asymptomatic proportion.
- Fit-for-purpose study designs, and improved reporting, are necessary for robust estimation of epidemiological characteristics from household studies and their interpretation.
- Ongoing assessment of the asymptomatic proportion of SARS-CoV-2 infection is critical to inform ongoing public health response options such as testing strategies to detect infections and isolation guidance for close contacts.

## Introduction

Sub-lineages of the SARS-CoV-2 Omicron variant of concern (VOC) continue to circulate globally and cause significant waves of transmission in the context of hybrid immunity from coronavirus disease 2019 (COVID-19) vaccination and infection.(1, 2) Although the Omicron-variant is associated with reduced disease severity relative to previous variants, it can still cause serious disease due to its ability to evade existing immunity.(3, 4)

Studies have shown that the viral load in the upper respiratory tracts of asymptomatic infected persons is comparable to that of symptomatic individuals, thus these individuals potentially contribute to onward transmission.(5-7) Asymptomatic individuals may be less likely to be indicated for or willing to test for infection, and rapid antigen tests (RATs) have been shown to have reduced diagnostic sensitivity in asymptomatic persons.(8-10) Further, individuals with asymptomatic infections may be less likely to practice social or physical distancing measures — due to not knowing they are infected and potentially infectious — and thus may contribute to the spread of infection in the general population.(11)

The asymptomatic proportion among SARS-CoV-2 Omicron-positive individuals has previously been estimated in two systematic reviews to be 32.4% (95% CI: 25.3–39.5%, *I*^*2*^ =97.7%) and 25.5% (95% CI: 17.0–38.2%, *I*^*2*^ =100%.(12, 13) Both reviews collated and synthesised data from various study designs, including cross-sectional studies that assessed symptom status at a single time point. Cross-sectional studies may lead to incorrect classification of presymptomatic individuals as asymptomatic — resulting in a biased estimate of the asymptomatic proportion — and could subsequently contribute to the high degree of observed heterogeneity when combined with estimates from sufficient study designs. Inclusion of such studies in a pooled estimate may lead to an overestimate of the asymptomatic proportion. Such evidence used for informing policy could lead to suboptimal testing of asymptomatic close contacts.

In 2020, the World Health Organization (WHO) developed the Unity Studies Early Investigation Protocols, to generate high-quality data to inform actions at the beginning of the COVID-19 pandemic.(14) One of the Unity protocols — for household transmission investigations (HHTIs) — provides a methodology for the systematic recruitment and longitudinal follow up of laboratory-confirmed SARS-CoV-2 cases and their household contacts, and collection of clinical, virological and serological data.(15) Systematic diagnostic testing and symptom data collection are needed to accurately ascertain infection events necessary to estimate the asymptomatic proportion of SARS-CoV-2 infection.

Our systematic review aimed to collate and synthesise the proportion of asymptomatic infections amongst household contacts of SARS-CoV-2 Omicron-variant positive cases, reported in studies aligned with the WHO Unity HHTI protocol. More specifically, we aimed to: identify and describe the implementation of HHTIs in time and place during SARS-CoV-2 Omicron-variant outbreaks; assess the methodological quality of included WHO-aligned HHTIs; calculate a pooled estimate of the asymptomatic proportion of SARS-CoV-2 Omicron-variant infections amongst household contacts, if appropriate, and; explore sources of heterogeneity in the included HHTIs.

## Methods

The systematic review protocol was registered on PROSPERO (CRD42022378648) and was conducted according to the Preferred Reporting Items for Systematic Reviews and Meta-Analyses (PRISMA) reporting guidelines.(16)

### Definitions

Asymptomatic SARS-CoV-2 infections were defined as infections confirmed through an appropriate diagnostic test — e.g., reverse transcriptase polymerase chain reaction (RT-PCR), or rapid antigen test (RAT) — where the individual experienced no symptoms consistent with the clinical case definition of COVID-19 (as defined by included studies in Supplementary Table 1). The asymptomatic proportion was defined as the number of asymptomatic secondary cases amongst all reported secondary cases in households.

**Table 1.** Summary characteristics of studies included in the systematic review of the asymptomatic proportion of SARS-CoV-2 Omicron-variant infections in households.

| Author, Year (ref) | Country | Relevant study period | Case ascertainment methods | Household contact testing strategy | Total number of household secondary cases | Total number of asymptomatic household secondary cases |
| --- | --- | --- | --- | --- | --- | --- |
| Smith-Jeffcoat et al., 2022 (21) | United States of America | 30 November 2021 – 20 December 2021 | RT-PCR and RAT | Unclear<sup>#</sup> | 6 | 0 |
| Bendall et al., 2022 (22) | United States of America | 1 November 2021<sup>+</sup> – 19 January 2022 | RT-PCR | 1) 'MHome' cohort - Days 0, 5, and 10 after enrolment for all participating household members.<br>2) 'HIVE' cohort - Days 0, 5, and 10 after the index case diagnosis | 23 | 11 |
| Ji et al., 2023 (23) | United States of America | November 2021 – March 2022 | RAT once daily upon enrolment <u>or</u> RT-PCR | Daily upon enrolment | 37 | 3 |

<sup>#</sup> Reported median symptom duration of 13 days among positive household contacts implies longitudinal data collection.
<sup>+</sup> Note that this study commenced on November 18, 2020. The reported dates relate to the identification of Omicron-variant infections, as relevant to this study.

Note that we include all cases in our estimate of the asymptomatic proportion besides the case that triggered recruitment to the study (i.e., the index case) — this was to avoid the potential bias that would result from a higher propensity for symptoms amongst the index cases.

### Search strategy

MEDLINE, EMBASE and Web of Science databases were searched to identify articles published between 1 November 2021 and 22 August 2023. We searched combinations of COVID-19, asymptomatic, household contacts, and Omicron (including Pango lineages BA.1, BA.2, BA.4 and BA.5). The medRxiv and bioRxiv preprint servers were also searched using the same search criteria. The detailed search strategies can be found in Supplementary Section S.1.

### Eligibility criteria

We included any published (peer-reviewed) or preprint article aligned with the WHO Unity HHTI protocol, involving five or more households, where household contacts were systematically tested for SARS-CoV-2 using an appropriate diagnostic test and had sufficient symptom data collected at more than one time point (i.e., not including cross-sectional studies). Only articles published in English were included.

Studies must have reported the proportion of asymptomatic or symptomatic infections amongst household contacts exposed to an index case of the Omicron-variant with a measure of uncertainty (e.g., confidence interval) or provided sufficient data to calculate these parameters.

### Screening and article selection

Records were imported into Covidence for de-duplication, storage, screening, and data extraction.(17) Records were screened by title and abstract by two independent reviewers (NS, AM) blinded to each other’s assessments, and a third independent reviewer (VS) resolved any conflicts. The same methods were applied to the full text screening.

### Data extraction

The following data fields were extracted using a structured and piloted form: investigation timing and duration of follow-up; definition of “household”; definition of “asymptomatic infection”; secondary case ascertainment methods; symptom data collection methods, and the number of index cases, households, household contacts, secondary cases, and asymptomatic secondary cases.

Where the reported estimates or definitions were unclear or not provided, study corresponding authors were contacted to request clarification or additional information. Investigations were excluded if authors did not respond after two email attempts over a four-week period.

### Methodological quality assessment

A critical appraisal tool for HHTIs was applied to the included investigations to assess their methodological quality.(18) Two independent reviewers (NS, AM) applied the critical appraisal tool and responses for each question were recorded as yes, no, or unclear. Each investigation was then determined to have a high, moderate, or low overall risk of bias.

### Data synthesis and statistical analysis

Estimates of the SARS-CoV-2 asymptomatic proportion – and associated 95% confidence intervals – were extracted from included articles or calculated from the raw data. Data cleaning and collation was performed using R version 4.0.(19)

## Results

Figure 1 summarises the literature search and screening process. We identified 8,368 records from the research databases and preprint servers, of which 3,770 were duplicates. Full text review was undertaken on 98 records. Three studies met our inclusion criteria and were retained for data extraction. Details of the reasons for exclusion are included in Figure 1.

**Figure 1.**
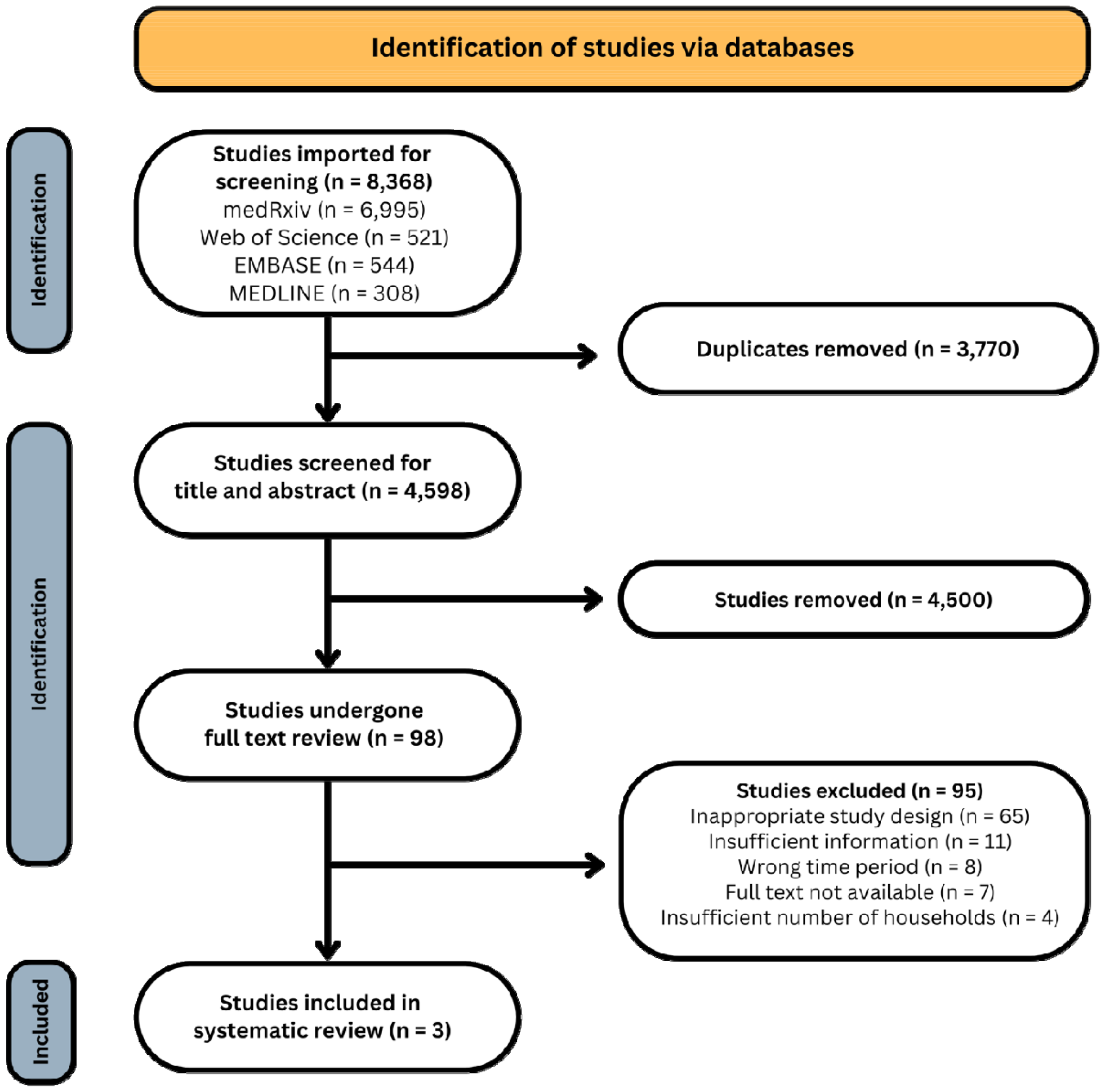
PRISMA flow chart.

The three included studies were all based in the United States of America and were conducted from November 2021 to March 2022. (20-22)

The estimates of the asymptomatic proportion from these three studies vary substantially, with point estimates of 0%, 6.7% and 47.8% (Figure 2). Further, these estimates were based on small sample sizes – the largest including 45 secondary cases. As a result, there is substantial uncertainty in each estimate, with the confidence intervals spanning 0–69.4%. Given the small number of included studies, we do not report a pooled estimate. Further, we have not calculated the *I*^*2*^ metric, due to its unreliability when the number of studies included is small.(20)

**Figure 2.**
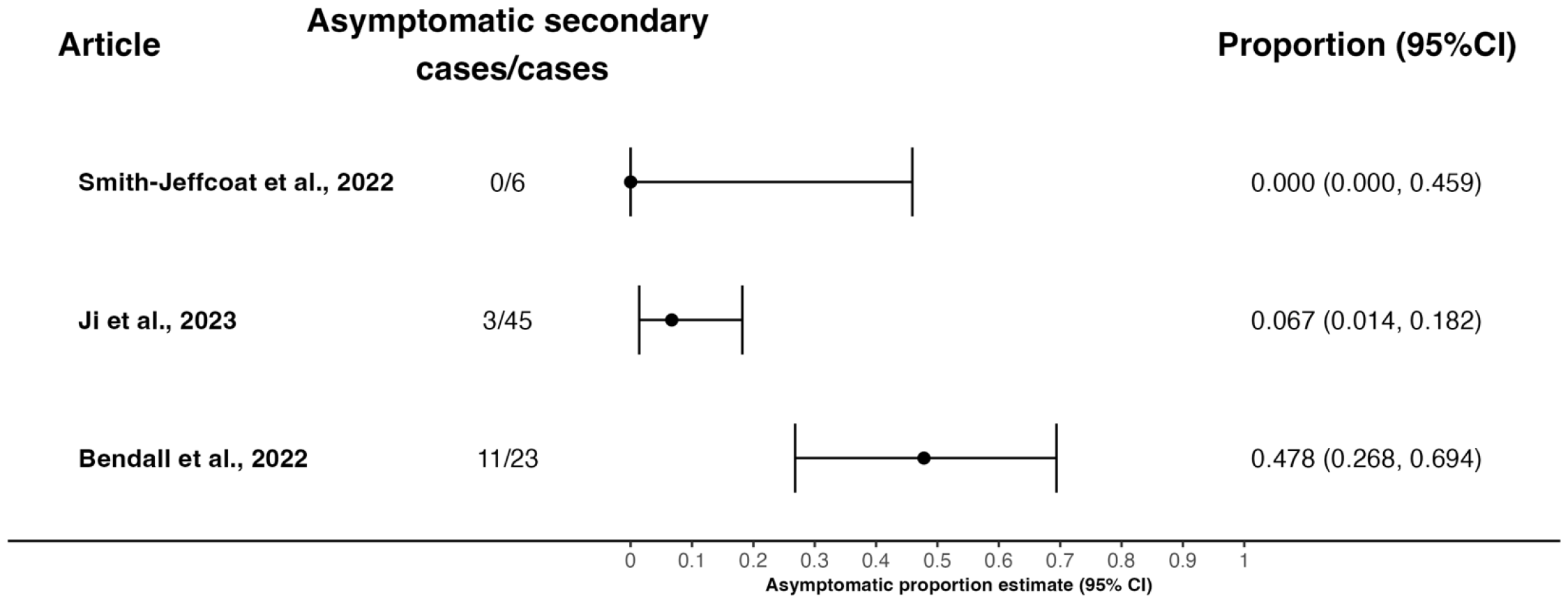
Forest plot of the asymptomatic proportion among household secondary cases. The estimated asymptomatic proportion and 95% confidence interval (CI) are shown on the right. Note: Smith-Jeffcoat et al., has a 97.5% confidence CI as zero asymptomatic infection events were observed.

### Smith-Jeffcoat et al. (2022)

Smith-Jeffcoat et al., (2022) recruited index cases from attendees at a convention in New York City, USA, which was held between 19 and 21 November 2021.(21) SARS-CoV-2 infections in attendees were identified using a combination of RT-PCR and RATs. Although the exact duration of follow-up was unclear, the investigators stated that median symptom duration was 13 days, implying that participants were followed for more than 13 days.

In total, 16 index cases (households) were identified who had 20 household contacts. Of these, six became secondary cases — all six cases were symptomatic during their infection.

Most participants — 100% (16/16) of index cases and 95% (19/20) of household contacts — completed their primary COVID-19 vaccine series more than 14 days prior to the study. Additionally, 38% (6/16) of index cases and 53% (10/19) of household contacts had received a booster dose.

### Bendall et al. (2022)

Bendall et al., (2022) recruited households in South-East Michigan, USA between November 1 2021 and January 19 2022.(22) Household were recruited into two cohorts, known as ‘MHome’ and ‘HIVE’. Participants in both cohorts were tested using RT-PCR at three timepoints during the investigation on Days 0, 5, and 10 after enrolment (‘MHome’) or after the index case diagnosis (‘HIVE’).

In total, 14 index cases (households) were identified, with 24 household contacts. Twenty-three household contacts became secondary cases, 11 of which remained asymptomatic after follow-up.

No vaccination information of householders was provided, however, 54.4% of the population in Michigan had completed their primary series of COVID-19 vaccinations by November 2021.(24)

### Ji et al. (2023)

Ji et al., (2023) recruited households through the California Institute of Technology in California, USA, between November 2021 and March 2022.(23) Only household members aged 6 years and older were included. Household contacts were tested daily using RATs as well as providing symptom information. 96% of participants were screened for at least 5 days and 53% were enrolled for at least 9 days.

In total, 28 index cases (households) were identified. An additional five households were recruited, where the infecting SARS-CoV-2 variant was inferred to be Omicron based on local predominance. The 33 index cases were associated with 130 household contacts (of which 109 related to the Omicron-variant confirmed households). Forty-five household contacts of Omicron-variant confirmed cases became secondary cases, and out of 37 secondary cases with complete symptom data, three remained asymptomatic until the end of follow-up.

31.3% (51/163) of participants (Omicron confirmed and inferred) received their primary COVID-19 vaccine series more than 7 days prior to the study, with 44.2% (72/163) receiving an additional booster dose. A further 3.7% (6/163) of participants were either unvaccinated or partially vaccinated (one dose), and the vaccination status was unknown for 20.9% (34/163) of participants. The incomplete symptom and vaccination status data was attributed to incomplete household recruitment, such that enrolled householders reported on the symptom status of their household members who chose not to directly participate.

## Discussion

This is the first systematic review investigating the asymptomatic proportion of SARS-CoV-2 infections in Unity-aligned HHTI studies. We identified three studies conducted in the United States from late-2021 to early-2022, with sufficient longitudinal follow-up and specimen sampling from household contacts. Effective control of an infectious disease requires identification and appropriate management of infectious individuals to prevent transmission. Infectious individuals not presenting with symptoms or meeting the clinical criteria of a case definition are typically harder to identify, and thus manage. As such, quantifying the prevalence of asymptomatic infections is critical to inform effective management strategies that do not rely on the presence of symptoms alone.(25)

The point estimates of the asymptomatic proportion ranged from 0–47.8%. Previously published systematic reviews produced pooled estimates of the asymptomatic proportion among SARS-CoV-2 Omicron-positive individuals of 32.4% (95% CI: 25.3– 39.5%) and 25.5% (95% CI: 17.0–38.2%), despite noting high levels of heterogeneity. The point estimates of included studies ranged from 1–92%, *I*^*2*^ = 97.7% and 4–40%, *I*^*2*^ = 100%.(12, 13) As noted above, we do not report a pooled estimate or *I*^*2*^ metric.

Two earlier systematic reviews of the ancestral SARS-CoV-2 strain, both published in 2020, by *Buitrago-Garcia et al. (26)* and *Byambasuren et al. (27)*, included 79 and 13 outbreak or contact tracing studies respectively, to produce a pooled estimate of the asymptomatic proportion. The included studies in each review were highly heterogenous, with estimates for the asymptomatic proportion ranging from 1–92% (pooled estimate of 19%, 95% CI: 17–25%), and from 4–40% (pooled estimate of 17%, 95% CI: 14–20%).(26, 27) Although cross-sectional studies were excluded from evidence synthesis in these reviews, the extent of heterogeneity as measured by *I*^*2*^ remained high *(84% in (27))*.

It is crucial to understand and contextualise the differences between studies prior to pooling estimates, to ensure each study is providing quality information towards the same quantity. This includes differences in study design (e.g., frequency and method of testing and symptom data collection), as well as any differences in population-level susceptibility (e.g., age-specific differences, or protection acquired through COVID-19 vaccination and/or SARS-CoV-2 infection), or public health and social measures (e.g., physical distancing or use of PPE and testing accessibility), that may influence detection of infection and extent of clinical disease. It may be that these differences contributed to the heterogeneity we observed in our three included studies.

We focused on household studies as a subset of the literature and thus had fewer studies suitable for inclusion, where previous reviews covered a broader range of study designs. The abundance of literature early in the pandy emic was likely due to the global need to accurately characterise the emerging virus as early as possible. Further, the expansion of testing and contact tracing capacities in 2020 and 2021 (prior to widespread COVID-19 vaccination) would have enabled or improved capacity for conducting studies with systematic testing and follow-up of close contacts, including HHTIs.

Subsequently, public health strategies shifted to impose less stringent PHSMs than had been implemented to-date, relying instead on effective vaccines against disease to reduce the burden of COVID-19. The substantial increase in caseloads corresponding to the spread of the Omicron-variant overwhelmed public health systems,(28) reducing the ability to implement the level of detailed contact-tracing required to generate data for ongoing characterisation of COVID-19, including the asymptomatic proportion. This shift in priorities was reflected in the literature during our full text review. Many excluded records did not estimate or report the asymptomatic proportion or have sufficient follow-up or testing strategies to reliably do so. Instead, they often focussed on the effect of vaccination on SARS-CoV-2 transmission dynamics, including estimates of secondary attack rates and vaccine effectiveness.

More generally, during the early stages of the pandemic, journals helped to expedite COVID-19 research to rapidly and widely disseminate information needed to address the global public health crisis. Our search strategy included articles published to 23 October 2023. Despite nearly being two years since the emergence of Omicron, we only found three relevant articles in the literature, which were all conducted in late-2021 to early-2022. In addition to changes in public health priorities and capacity to conduct HHTIs, changes in the dissemination of COVID-19 studies in scientific journals may have also delayed the availability of other, relevant studies at the time of this review. We tried to account for delays in publication as the pandemic progressed by searching the medRxiv and bioRxiv pre-print servers, which were commonly used to rapidly disseminate articles prior to publication.

While we targeted Unity-aligned studies in our review with the aim of improving comparability of studies, there were still differences present in the design and implementation of included studies. For example, age-specific severity and duration of follow-up are both important aspects for measuring and estimating severity indicators, including the asymptomatic fraction. Age-dependent severity has been documented extensively for SARS-CoV-2 infections since the ancestral variant.(29) However, reporting of severity indicators such as the asymptomatic fraction, are not routinely adjusted by age – including in the studies in this review. Further, one study did not include children under 6 years of age. The absence of age-adjusted information — and other underlying characteristics and/or risk factors — makes it challenging to explore whether differences in cohorts across studies are substantial contributors to the heterogeneity in reported estimates of the asymptomatic fraction. The follow-up duration differed across the three studies — between 5 and at least 13-days post-recruitment of the index case — as well as the frequency of testing and collection of symptom status. The incubation period for the Omicron-variant of SARS-CoV-2 has since been estimated in many studies (30-32) — one estimated the median incubation period to be 3.8 days (95%CrI 3.5–4.1).(30) If this distribution was observed in participants in our study, it would be expected that only 82.5% (95%CI 75.5–88.1) of secondary cases would present with symptoms by day 5 were they infected at recruitment (and less if infected thereafter). Further, while we defined inclusion based on individuals being classified as secondary cases, the literature often did not distinguish secondary and subsequent cases (or unrelated cases) in transmission chains. As a result, there is an increased likelihood that our included studies still incorrectly classified individuals as pre-symptomatic. The design of transmission studies (including HHTIs) to infer epidemiological characteristics of a pathogen should consider the range of possible values each quantity could take. In the case of the asymptomatic fraction, study design should consider the generation interval distribution and incubation period distribution to ensure that participants that are infected (possibly after recruitment given ongoing exposure to cases) are followed for sufficiently long to accurately record their symptom status throughout their infection. Where this information is unknown at the time, a conservative approach to defining a sufficient length of follow up should be taken to avoid resources wastage where quantities cannot be estimated due to an inappropriate study design.

In light of lessons learned through the COVID-19 pandemic thus far, WHO have released updated Unity protocols for influenza and pan-respiratory viruses.(33, 34) The sampling schedules therein correlate with biological and epidemiological quantities of each exemplar pathogen to guide appropriate data and specimen collection to inform classification of subsequent cases. Modelling studies accounting for uncertainty in these quantities in different pathogen scenarios should be undertaken to inform optimal sampling schedules.(35-37)

The heterogeneous evidence for the asymptomatic proportion used in existing systematic reviews for ancestral SARS-CoV-2 and Omicron suggests that study designs still need to be standardised for better implementation and reporting across different settings and populations. This is a consistent message with a recent review of the household secondary attack rate,(38) where similar limitations in study design and reporting were identified, which motivated the development of reporting guidelines and updates to the WHO protocols.(18, 33, 34)

Although the Unity Studies were motivated to produce early generation of evidence for COVID-19, HHTIs should be used to support ongoing monitoring of epidemiological quantities as the COVID-19 pandemic evolves, e.g., during the emergence of Omicron subvariants or new VOCs. Ongoing assessment of these quantities is critical so that public health response options such as testing strategies to detect infections and isolation guidance for close contacts, can be adjusted accordingly.

## Conclusion

HHTIs remain a valuable tool to collect data and collate information on key clinical and epidemiological data of COVID-19, especially given the continued evolution of SARS-CoV-2. The limited number of quality studies that we identified highlights the need for improved preparedness and response capabilities to facilitate robust HHTI implementation, analysis and reporting, to better inform national, regional and global risk assessments and policy making.

## Data Availability

The data underlying this article are available in the article and in its online supplementary material.

## Ethics approval

Not required

## Supplementary Data

Supplementary data are available at *IJE* online.

## Author contributions

The authors confirm contribution to the paper as follows: study conceptualisation and methodology – all; investigation — NDJS, AM, VS; data curation — NDJS, AM; visualization — NDJS, AM, DJP; supervision — AM, VS, JPVC, DJP; writing – original draft — NDJS, AM, DJP. All authors reviewed and edited the draft manuscript and approved the final version for publication.

