## Supplementary Material for "The asymptomatic proportion of SARS-CoV-2 Omicron-variant infections in households: A systematic review"

**Supplementary information**

**S.1 | Detailed Search Strategy**

1. Medline, EMBASE, Web of Science and Scopus — searched 22 August 2023

(COVID-19 or COVID19 or SARS-CoV-2 or coronavirus)

AND

(asymp* or symp* or sever*)

AND

(household* or famil* or contact* or residen* or dwelling*)

AND

(Omicron or "B.1.1.529" or "BA.1*" or "BA.2*" or "BA.3*" or "BA.4*" or "BA.5*" or "BA1*" or "BA2*" or "BA3*" or "BA4*" or "BA5*")

NOT

(mouse* or mice* or hamster* or ferret* or "animal model*" or KAP* or "mental health*" or "long term care facil* or "nursing home*" or "aged care*")

2. medRxiv and bioRxiv — searched between 1 November 2021 and 22 August 2023

Advanced Search of (abstracts or titles)

Covid “asymptomatic”

Covid “symptomatic”

Covid “severe”

Covid “severity”

Covid “household”

Covid “family”

Covid “contact”

Covid “resident”

Covid “Omicron”

Covid “B.1.1.529”

Covid “BA.5”

Covid “BA.4”

Covid “BA.3”

Covid “BA.2”

Covid “BA.1”

**S.2** | **Detailed questions used for methodological quality assessment**

This systematic review used a bespoke critical appraisal tool designed specifically to assess the methodological quality of HHTIs.(1) We used all questions to guide an overall assessment of the risk of bias in relation to our primary objective of estimating the asymptomatic proportion among household contacts of laboratory confirmed Omicron-variant cases.

The questions detailed in the HHTI critical appraisal tool used to assess these studies are as follows:

| **Q1** | Was the timing of recruitment and data collection appropriate to achieve the objectives of the investigation? |
| --- | --- |
| **Q2** | Was the method for index case ascertainment appropriate? |
| **Q3** | Was a definition of 'household' provided? |
| **Q4** | Were all eligible cases and all householders enrolled into the investigation? |
| **Q5** | Were subsequent cases identified and ascertained using appropriate methods? |
| **Q6a** | Was the duration of follow-up sufficient to measure outcomes? |
| **Q6b** | Did all participants remain part of the 'household' for the duration of the investigation? |
| **Q7a** | Were steps taken to ensure that householders were susceptible at the time of enrolment? |
| **Q7b** | Were steps taken to ensure that subsequent infections were due to exposure within the household? |
| **Q8** | Are the analytic methods appropriate given the study context and design? |
| **Q9** | Has loss-to-follow-up been appropriately accounted for in the estimated outcomes? |
| **Q10** | Has any missing data been appropriately accounted for in the estimated outcomes? |

**S.3 | Detailed characteristics of the included studies**

| Author, Year (ref) | Study period | Population description | Definition of household | Specific symptoms included in each paper consistent with COVID-19 | Definition of asymptomatic infection | Definition of symptomatic infection |
| --- | --- | --- | --- | --- | --- | --- |
| Smith-Jeffcoat et al., 2022 (2) | November 30 2021 — December 20 2021 | Individuals who attended a New York City convention (who were interviewed after testing positive) and their household contacts. | An individual who slept in the same household at least once (and was not an attendee at the convention exposure event) in the two weeks following the attendees' return from the convention. | Not specifically provided – nasal congestion, fatigue, cough, runny nose and sore throat were commonly reported symptoms. | An infection confirmed through testing where the individual has no symptoms consistent with COVID-19 throughout the duration of their whole infection. | Not provided |
| Bendall et al., 2022 (3) | November 18 2020 — January 19 2022 | Households were enrolled in two household cohorts - 'MHome' cohort and 'HIVE' cohort.   'MHome' cohort - case ascertained household cohort in which households are recruited following identification of an index case who meets a case definition for COVID-like illness and is positive for SARS-CoV-2 by clinical testing.   'HIVE' (household influenza vaccine evaluation study) cohort - prospective household cohort with year-round surveillance for symptomatic acute respiratory illness, with index case and all household members included. | MHome' cohort - not stated but assumed the same - "households are recruited following identification of an index case who meets a case definition of COVID-like illness and is positive for SARS-CoV-2 by clinical testing"  'HIVE' cohort - households with 1 or more individuals positive for SARS-CoV-2 | In the HIVE cohort, eligible illness was defined as two or more of: cough, nasal congestion, sore throat, chills, fever/feverish, body aches, or headache (for participants 3 years & older) or two or more of cough, runny nose/nasal congestion, fever/feverish, fussiness/irritability, decreased appetite, trouble breathing, or fatigue (for participants under 3 years old).   In the MHome cohort, index enrollees meeting the case definition (at least one the following: cough, difficulty breathing, or shortness of breath; or at least two of the following: fever, chills, rigors, myalgia, headache, sore throat, new loss of smell or taste). | Not provided | Two or more of cough, nasal congestion, sore throat, chills, fever/feverish, body aches, or headache (for participants 3 years & older) or two or more of cough, runny nose/nasal congestion, fever/feverish, fussiness/irritability, decreased appetite, trouble breathing, or fatigue (for participants under 3 years old) |
| Ji et al., 2023 (4) | November 2021 - March 2022 | Households were included in this analysis if laboratory testing confirmed that at least one household member was acutely infected with SARS-CoV-2 and more than a third of reported household members were enrolled | Not provided | Defined according to CDC https://www.cdc.gov/coronavirus/2019-ncov/symptoms-testing/symptoms.html | Infection where individual reported no COVID-19 symptoms prior to enrolment nor at any RT-qPCR positive timepoint | Not provided |

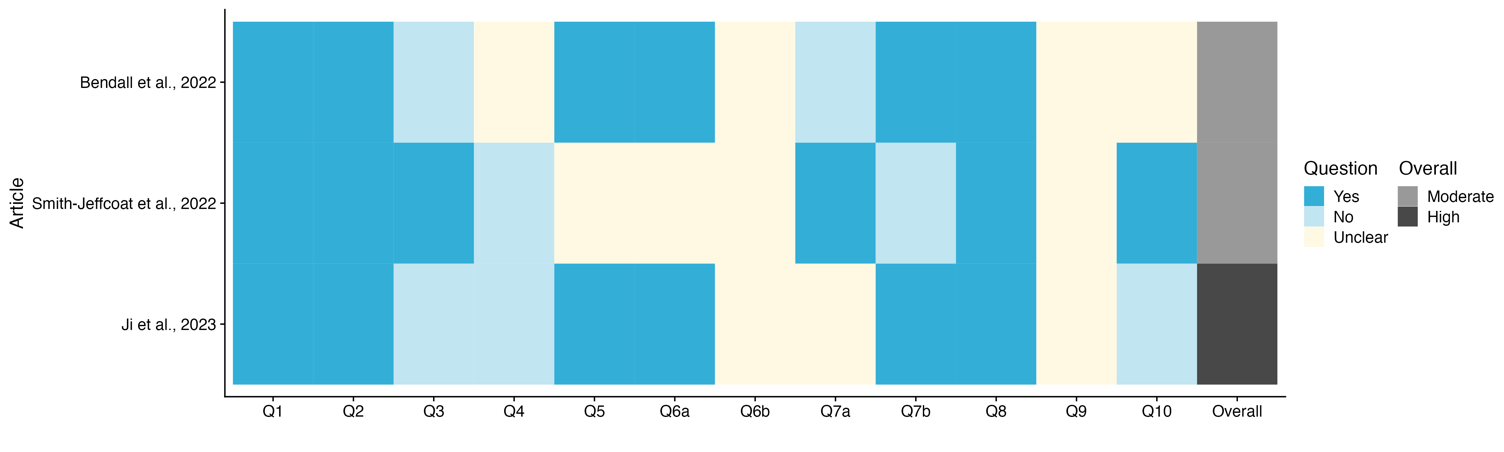
**S.4 | Results of the critical appraisal tool**

**Supplementary Figure 1.** *Results of the critical appraisal tool as applied to articles that reported the asymptomatic proportion among household contacts. Colours for Questions 1–10 indicate whether each was addressed in the investigation (dark blue) or not (cream), or instances where there was insufficient detail available to assess (light blue). An overall rating of the risk of bias is provided in the far-right column, with investigations rated Medium (grey) or High (grey).*

**S.5 | References**

1.	Price DJ, Spirkoska V, Marcato AJ, Meagher N, Fielding JE, Karahalios A, et al. Household transmission investigation: Design, reporting and critical appraisal. Influenza and Other Respiratory Viruses. 2023;17(6):e13165.

2.	Smith-Jeffcoat S, Pomeroy M, Sleweon S, Sami S, Ricaldi J, Gebru Y, et al. Multistate Outbreak of SARS-CoV-2 B.1.1.529 (Omicron) Variant Infections Among Persons in a Social Network Attending a Convention — New York City, November 18–December 20, 2021. MMWR Morbidity and Mortality Weekly Report. 2022;71:238-42.

3.	Bendall EE, Callear AP, Getz A, Goforth K, Edwards D, Monto AS, et al. Rapid transmission and tight bottlenecks constrain the evolution of highly transmissible SARS-CoV-2 variants. Nat Commun. 2023;14(1):272.

4.	Ji J, Viloria Winnett A, Shelby N, Reyes JA, Schlenker NW, Davich H, et al. Index cases first identified by nasal-swab rapid COVID-19 tests had more transmission to household contacts than cases identified by other test types. PLOS ONE. 2023;18(10):e0292389.
